# A multi-level evidence approach to evaluate training, performance, and system-level impact of Hamdard Force: a community mental health intervention

**DOI:** 10.64898/2026.09.29.26363287

**Authors:** Asma Humayun, Noor ul Ain Muneeb, Asma Nisa, Ayesha Aziz

## Abstract

**Background:** There is limited evidence to demonstrate the impact of training community mental health workers beyond knowledge acquisition. This study evaluated the training, workforce performance, and system-level outcomes of the Hamdard Force, a community mental health intervention, implemented in two districts of Khyber Pakhtunkhwa, Pakistan, as part of a multi-tiered, digital MHPSS service over 8 months.

**Methods:** We followed a multi-level evidence approach, guided by the New World Kirkpatrick Model, using a mixed-methods design that combined quantitative and qualitative indicators. We assessed participants’ reactions, learning, behavioral outcomes, system-level outcomes of the intervention, and conducted Focus Group Discussions to identify contextual factors that influenced implementation.

**Results:** At Tier 1, a total of 403 community workers were trained, who reported satisfaction with the training content and relevance, and demonstrated significant improvement in knowledge post-training. Less than half of the trained workforce remained active in the field and referred people with mental healthcare needs under supervision. A total of 855 people were triaged by clinical psychologists at Tier 2 of the service model, and 549 cases met the criteria of being valid. The results of detailed clinical assessment and management demonstrated encouraging outcomes regarding the effectiveness of the intervention, both at the individual case and at the system-level. The Focus Group Discussions revealed the process of supervision, organizational support and digital tools of learning were significant drivers of the intervention. Challenges with internet connectivity, limited digital literacy, and persistent mental health stigma were identified as implementation barriers.

**Conclusion:** Overall, the integrated findings suggest that the intervention helped to build the capacity of the workforce, facilitated their performance through supervision, provided services through a task-sharing approach and strengthened referral pathways. These findings underscore the importance of implementation-focused evaluations for scaling up community mental healthcare in low-and middle-income countries.

## Introduction

Mental disorders continue to be among the top ten leading causes of global burden of disease and account for a substantial share of years lived with disability (Global Burden of Disease, 2022). The treatment gap for mental disorders is significantly high across low- and middle-income countries (LMICs), even for common conditions like depression, where only 3% of people receive minimally adequate treatment, compared to 23% in high-income settings (Moitra et al., 2022). Limited investment in public mental health and a significant dearth of trained mental health professionals, especially in rural and underserved areas, are major barriers to addressing mental health needs (World Health Organization, 2022). In a meta-regression analysis of the prevalence of common mental disorders across eight countries in South Asia, Pakistan showed the highest burden of mental illness, significantly exceeding Bangladesh, the reference country (Naveed et al., 2020).

The World Health Organization strongly recommends strengthening community-based mental healthcare to address the existing treatment gap in LMICs (World Health Organization, 2021). There are reports of many well-designed and scalable capacity-building initiatives that were either not integrated into national health systems or not sustained (Patel et al., 2018; Upadhyay et al., 2023). As Pakistan continues to face recurring humanitarian and climate-related challenges, the need to implement scalable interventions to build the capacity of community workers is critical. This workforce can act as frontline responders by identifying persons with needs early, providing them basic psychosocial support, and facilitating timely referral to appropriate services, particularly during humanitarian emergencies when access to mental health and psychosocial support is disrupted (Bolton et al., 2023; Patel et al., 2018; World Health Organization, 2021). However, most community interventions focus on improvement in knowledge and skills through short-term capacity-building workshops, which might not guarantee meaningful outcomes (Fox et al., 2012; Shultz & Forbes, 2014).

In 2021, the Ministry of Planning, Development & Special Initiatives (MoPD&SI) developed a multi-tiered, digital mental health and psychosocial support (MHPSS) service model, which is evidence-based and scalable. At Tier 1 of this model, a community mental health intervention, known as the Hamdard Force (HF), was developed to build the capacity of the community mental health workforce (Humayun et al., 2026). The training resources were adapted from Psychological First Aid (World Health Organization, 2011) and delivered through digital tools, including the MHPSS web portal^1^. A Learning Management System (LMS) for online training and the HF mobile application to receive data from the community workers are integrated with the portal. A key feature of the MHPSS model is to develop a mental health workforce: clinical psychologists (CPs) at Tier 2, primary care physicians at Tier 3, and specialists at Tier 4. The overall model is designed to develop a stepped care pathway to address existing treatment gaps (MoPD&SI, 2025).

The Hamdard Force intervention was piloted in Haripur and Kohat (two districts in the province of Khyber Pakhtunkhwa) from October 2025 to June 2026. The objectives of the current study were to 1) evaluate the training and performance outcomes of the pilot implementation of Hamdard Force intervention to identify, support and refer people with mental health needs; 2) assess system-level impact of the Hamdard Force intervention during the pilot period; and 3) explore the drivers and barriers that influenced the process of implementation, with the ultimate aim of scaling up the intervention across the province effectively.

## Method

### Phase 1: Recruitment and registration

We sought nominations for HF through intersectoral collaboration with governmental line departments (including health and education) and non-governmental organizations. Our inclusion criteria required at least 12 years of school education, experience in community engagement, effective communication skills, willingness to seek training and supervision using the program’s digital tools, and interest in supporting people with mental health conditions in the community. We also requested that both genders be equally represented in the nominations.

We employed purposive sampling and aimed for a cohort of 400 participants, but over-recruited 482 participants to account for expected attrition during training. A total of 440 participants registered, but only 403 participants completed the training.

The demographic characteristics of the trained workforce are presented in Table 1.

**Table 1.** Sociodemographic Characteristics of Trained Hamdard Force Members (N = 403)

| **S. No.** | **Demographic** | **Groups** | **n (%)** |
| --- | --- | --- | --- |
| 1 | Gender | Female | 309 (76.7) |
|  |  | Male | 94 (23.3) |
| 2 | Age (range 20–68) | 20–35 | 190 (47) |
|  |  | 36–50 | 159 (39.6) |
|  |  | >50 | 54 (13.4) |
| 3 | District | Haripur | 197 (48.9) |
|  |  | Kohat | 206 (51.1) |
| 4 | Education | 12 years of school education<sup>2</sup> | 161 (40.0) |
|  |  | Graduation | 129 (32.0) |
|  |  | Master's or above | 113 (28.0) |
| 5 | Occupation | Health sector | 235 (58.3) |
|  |  | Education sector | 83 (20.6) |
|  |  | Students and volunteers | 49 (12.2) |
|  |  | NGO/Other | 36 (8.9) |
| 6 | Years of work experience | <1 year | 52 (12.9) |
|  |  | 1–5 years | 71 (17.6) |

<sup>2</sup> Included: A Level / FSC / FA / ICS / ICOM / Intermediate diploma.

|  |  |  |  |
| --- | --- | --- | --- |
|  |  | >5 years | 280 (69.5) |

### Phase 2. Training

We conducted a total of 13 (half-day) training workshops in each district in October 2025. Each workshop was attended by 15-20 participants. The training was conducted through a blended approach where HF members were facilitated by on-site supervisors (CPs at Tier 2) to complete the course online and use the HF mobile application to refer cases. Each workshop was supervised by CPs who also assisted with technical troubleshooting.

The participants registered on the MHPSS web portal and logged into the LMS for self-directed HF courses (either in Urdu or in English). The courses were structured around 10 interactive modules, which comprised written content, audio narration, locally relevant illustrations, and quizzes to leverage the testing effect. The average time taken to complete the course was 2 hours. After completing the course, the HF members also received a copy of the printed HF guide in Urdu as a reference. After completing the course, the HF members downloaded the HF mobile application, which served as an interface between the HF members and the MHPSS portal. It is a simple, user-friendly application designed for remote areas with limited internet access, requires low bandwidth, and is usable offline. During the training, each HF member submitted a test case under supervision to practice the process of case entry. During fieldwork, they were required to obtain consent from a person with MHPSS needs in the community; enter their name, age, contact details, and briefly state the reason for referral. Where possible, they were also guided to complete the GHQ screening after obtaining consent.

### Phase 3: Evaluation of the training

We followed a multi-level evidence approach guided by the New World Kirkpatrick Model (NWKM) (Kirkpatrick & Kirkpatrick, 2016) to evaluate the training and performance of the HF intervention. The NWKM framework positions the training and performance outcomes within a results-oriented framework and has been applied to complex training programmes (Liao & Hsu, 2019). In addition, it acknowledges the role of ‘required drivers’ in supporting training outcomes, uses ‘leading indicators’ as early signals of broader impact, and explains the evaluation across all levels by building a coherent ‘Chain of Evidence’(Kirkpatrick & Kirkpatrick, 2016). Our study extended beyond participants’ reaction and learning outcomes to examine behavioral outcomes and broader system-level impact, and offered a chain of evidence across four levels to integrate community-based mental health intervention into the existing healthcare system.

### Level 1: Reaction

We used a mixed-methods design combining quantitative and qualitative feedback to assess the reaction of the participants to the training. The HF members submitted their quantitative feedback through a structured 7-item questionnaire using a three-point response scale: Good, Needs improvement, Poor; previously designed for the intervention (Humayun et al., 2025a). The questionnaire helps assess the course content, design, duration, use of illustrations and audio recordings, assessment quizzes, and language. The digital form (on the LMS) also included an open-ended question at the end for the participants to provide their comments.

### Level 2: Learning

The training data was obtained from the MHPSS web portal. 91.5% of the registered participants completed training, corresponding to an overall attrition of 8.5% from registration to training completion. Among those who completed training, nearly two-thirds selected the Urdu-language course and just under a third selected the English-language course. We assessed knowledge acquisition using a 20-item pre- and post-test already developed for evaluating HF training (Humayun et al., 2025a).

### Level 3: Behavior & field performance

Each HF member was assigned to a supervisor (a CP at Tier 2) and was part of a supervision group on WhatsApp. The supervisors assisted the HF members with technical and clinical aspects, encouraged them to identify and refer cases, monitored the referred cases on their dashboards, and provided feedback on their performance.

We evaluated the post-training behavior and field performance of the trained HF members after 8 months using quantitative and qualitative indicators as follows:

3.1 Number of active HF (HF members who referred at least one case).
3.2 Number of valid cases At Tier 2, the CPs assessed and managed the referred cases. Cases that could not be contacted; or were not interested; or did not have any MHPSS needs; or did not reside in the target districts were marked as ‘invalid’ and excluded from further analyses.
3.3 Assessment skills The CPs re-assessed the screening for psychological well-being reported by the HF members to evaluate the assessment skills of HF.
3.4 Supervisory appraisal
We developed an appraisal form on the dashboard of the supervisors to evaluate the performance of each HF member in the last month of the pilot implementation, considering level of participation; quality of documentation; accessibility and responsiveness; level of interest in seeking supervision. The overall appraisal was based on a three-point scale (3=above average, 2=average, 1=below average):

a. Above average indicated active participation, referred many cases, complete documentation, very accessible and responsive, and interested in seeking supervision when needed.
b. Average indicated participation, referred some cases, sometimes accessible and responsive, and limited interest in seeking supervision.
c. Below average indicated limited participation, referred a few cases only, limited accessibility and responsiveness, reported the case in a single word, and used stigmatizing language, with reduced interest in seeking supervision.
3.5 A multi-dimensional evaluation of the performance We triangulated the field performance indicators and supervisory appraisal to assess a multidimensional evaluation of the performance of active HF members.
3.6 Field performance as a test of recruitment criteria We conducted an analysis of characteristics of trained HF members as predictors of field performance to strengthen our recruitment criteria.

### Level 4: System-level impact

After measuring the performance of the HF members, we assessed the impact of their performance, reflecting the downstream, system-level impact of the outcomes of cases referred by the HF members. This was done by examining the outcomes of the referred cases, as assessed and managed by the CPs (at tier 2) using the following indicators:

4.1 Assessment

a. Psychosocial stressors
b. MHPSS needs
c. Duration of mental health conditions
d. Social support
4.2 Psychosocial interventions The CPs managed stress-related conditions, encouraging individuals to use MyCare+ mobile application for self-guided stress management. MyCare+ app is part of the MHPSS service model, hybrid in approach and designed as a scalable intervention for stress management (Humayun et al., 2020). The CPs, trained in implementing mhGAP-Humanitarian Intervention guidelines, provided psychosocial interventions for other mental health conditions as well.
4.3 Follow-up and referral pathway

a. No of calls We monitored the number of assessments, including follow-up calls made by the CPs to their assigned cases.
b. Further referral The CPs further referred individuals to the MHPSS team, mostly when a local psychiatrist was not available or inaccessible due to long distance or socioeconomic barriers; nearest primary care physicians (trained in implementing mhGAP-Humanitarian Intervention guidelines at Tier 3), particularly where pharmacological interventions were indicated); local psychiatric service; and other services.

### Implementation drivers and barriers

In addition to measuring the impact of our intervention, we conducted 4 Focus Group Discussions (FGDs) to identify drivers and barriers to recruitment, training, and performance of HF. Two FGDs were conducted with HF members (n= 20 each), one with their clinical supervisors (CPs) (n=13), and one with the MHPSS team providing overall technical support (n=5). Semi-structured questionnaires were employed to explore experiences related to selection and recruitment, delivering the training, digital accessibility, supervision, and referral of cases (added as supplementary material A). The sessions were audio-recorded, transcribed, and analyzed using thematic analysis.

Data source triangulation was used to integrate findings from different stakeholder groups to examine both shared and divergent perspectives, enabling a richer and more comprehensive understanding of participants’ experiences (Morgan, 2024).

## Results

### Level 1: Reaction

The results of participants’ feedback on HF training were positive across all components (see supplementary material B). They expressed their satisfaction with the course design, deeming the content relevant to their context and needs. The illustrations and audio recordings were found to be helpful in enhancing the understanding of the content. The language was considered simple, used in daily life rather than being text-heavy. The digital format was well-received by most members, while some reported longer completion times, particularly in Kohat, where internet connectivity issues affected the training experience. In some instances, digital literacy affected user experiences, as participants completed the course in 5 to 6 hours and found interactive assessment quizzes difficult to attempt, requiring improvement.

Consistent with the quantitative findings, qualitative feedback also indicated that participants found the training useful and reported improvements in their knowledge, confidence, and ability to provide support in community settings.

### Level 2: Learning

The post-training knowledge scores (M=12.52) were significantly higher than pre-training scores (10.48, p <0.001). No major difference was observed in knowledge assessment for English and Urdu courses, as there was a significant increase in assessment scores post-training in both courses.

### Level 3: Behavior & field performance

#### 3.1 Number of active HF

During the implementation, 160 HF members (39.7%) were classified as ‘active HF’, indicating that 60.3% of trained members did not become active in field referral. Among the active HF members, 123 referred more than one case. The recruitment, training, and performance outcomes are illustrated in Figure 1.

**Figure 1:**
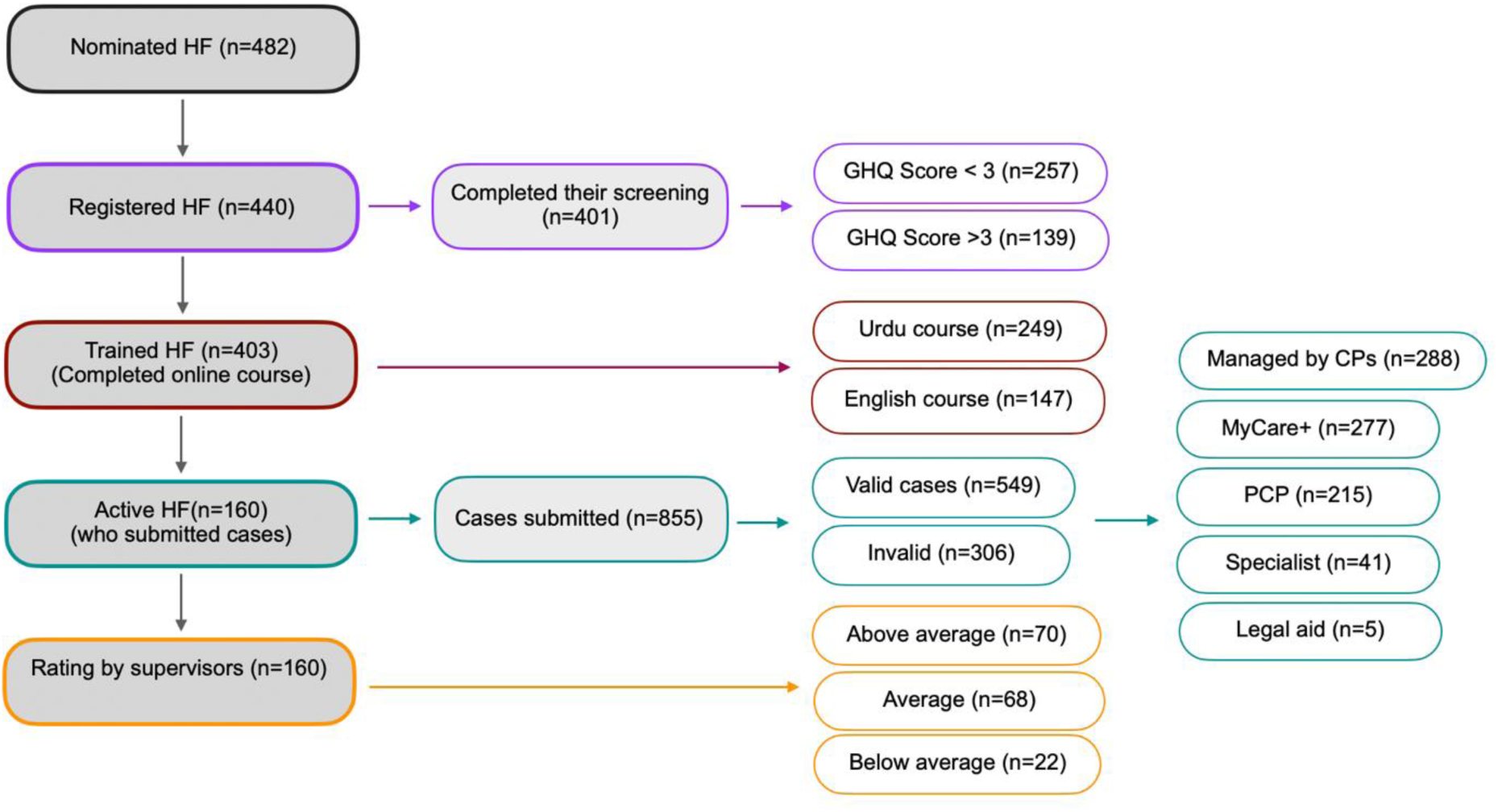
Flow diagram of Hamdard Force recruitment, training and performance outcomes.

#### 3.2 Number of referred valid cases (Quantitative performance indicator)

Out of a total of 855, 549 were valid cases referred from the community by the HF members. 391 (71.2%) cases were routine, while 158 (28.8%) were urgent referrals.

##### Demographics of referred cases

The age range of the service users ranged from 4 to 93 years. 65.5% of the referred cases were females. 423 cases (49.5%) were referred from Haripur, while 432 cases (50.5%) were referred from Kohat. 24.6% were students, while 19.3% were homemakers and 18.2% were employed. The majority of the service users were married (40.1%), while 35.5% were single. The demographic table is added as Supplementary material C.

#### 3.3 Quality of assessment

Table 2 illustrates comparisons of GHQ-12 scores assessed by HF members at initial screening with those recorded by CPs following reassessment. HF members tended to overclassify service users in the higher-distress category, whereas CPs classified a lower proportion of service users in the high-distress category and more in the low-distress category, indicating refinement of community-based screening during clinical review.

**Table 2:**
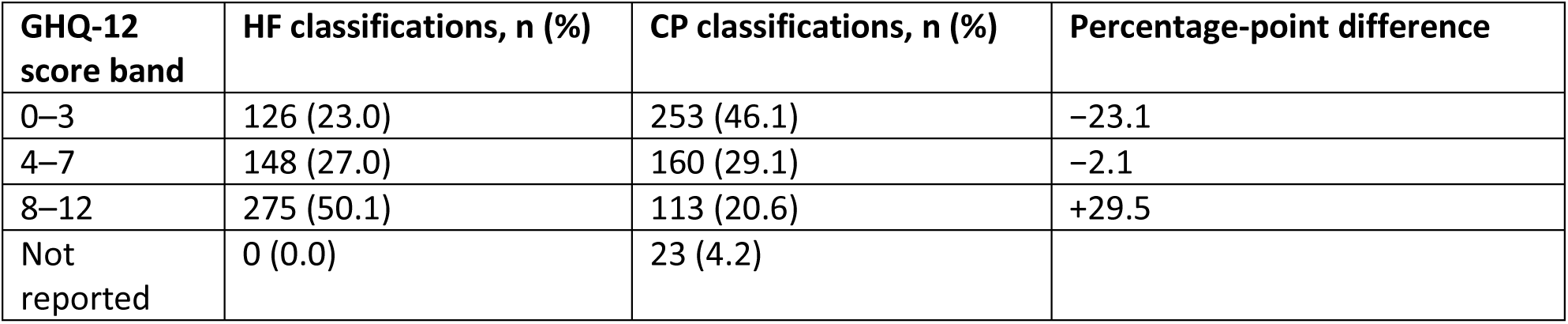
GHQ-12 scores by HF members and re-assessed by clinical psychologists.

|  |  |  |  |
| --- | --- | --- | --- |
| **GHQ-12 score band** | **HF classifications, n (%)** | **CP classifications, n (%)** | **Percentage-point difference** |
| 0–3 | 126 (23.0) | 253 (46.1) | –23.1 |
| 4–7 | 148 (27.0) | 160 (29.1) | –2.1 |
| 8–12 | 275 (50.1) | 113 (20.6) | +29.5 |
| Not reported | 0 (0.0) | 23 (4.2) |  |

#### 3.4 Supervisory appraisal

70 (43.8%) HF members were rated as above average, 68 (42.5%) as average, and 22 (13.8%) as below average (see Table 3).

**Table 3:** Supervisory appraisal vs number of cases referred.

| **Table 3: Supervisory appraisal vs number of cases referred** |  |  |  |
| --- | --- | --- | --- |
| **Supervisor rating** | **No of HF (n)** | **Valid cases** | **Median valid cases per HF** |
| Above average | 70 | 353 | 5 |
| Average | 68 | 151 | 1 |
| Below average | 22 | 45 | 1 |

**Note:** Median values are presented due to the skewed distribution of case counts.

**Table 4.** Participant characteristics predictive of being an active HF.

| **Table 4. Participant characteristics predictive of being an active HF** |  |  |  |  |  |
| --- | --- | --- | --- | --- | --- |
| **Variable** | **Category** | **Active HF, n/N (%)** | **Adjusted OR** | **95% CI** | **p-value** |
| **Age** | $> 35$ years | 58/213 (27.2) | — | — | — |
| | $\leq 35$ years | 102/190 (53.7) | 1.70 | 1.03–2.82 | .039 |
| **Education** | Intermediate | 39/161 (24.2) | 0.53 | 0.30–0.93 | .027 |
|  | Graduation | 65/129 (50.4) | — | — | — |
|  | Master's or above | 56/113 (49.6) | 1.49 | 0.86–2.59 | .160 |
| **Work experience** | <1 year | 38/52 (73.1) | 3.36 | 1.52–7.70 | .003 |
|  | 1–5 years | 34/71 (47.9) | — | — | — |
|  | >5 years | 88/280 (31.4) | 0.79 | 0.43–1.43 | .427 |

**Note.** *N* = number of HF members and *n* = number of active HF members in the corresponding category. Percentages represent active HF within each category. Reference categories were >35 years for age, Graduation for education, and 1–5 years for work experience. OR = odds ratio; CI = confidence interval.

#### 3.5 A multi-dimensional evaluation of the performance

A Spearman correlation analysis showed significant positive associations between supervisor ratings and the number of valid cases (Spearmen’s p= .536, p <.001). Higher supervisor ratings were associated with greater case reporting activity (Table 3).

A Kruskal–Wallis test showed a significant difference in valid case reporting across the three supervisor rating categories (χ² (2) = 50.22 p < 0.001). Bonferroni-adjusted post hoc comparisons showed that HF members rated as above average reported significantly more valid cases than those rated as average or below average. No significant difference was observed between the average and below-average categories.

Supervisor ratings did not differ significantly across gender, age group, district, education, occupation, work experience, or prior mental health training.

#### 3.6 Field performance as a test of recruitment criteria

We conducted a multivariable logistic regression analysis to determine whether being an active HF was independently associated with age, education, and work experience. HF members aged ≤35 years had significantly higher odds of being active than older members.

Graduate participants had significantly higher odds of being active than those with intermediate education. Whereas there was no significant difference when comparing graduates with those with higher education. Members with <1 year of work experience had significantly higher odds of being active than those with 1-5 years of experience. No significant difference was observed between members with >5 years and those with 1-5 years of experience.

Descriptive cross-tabulations between age and education showed that a higher proportion of younger HF members were active across all education categories.

Descriptive cross-tabulations between age and work experience (see supplementary material D) showed that younger HF members performed better than older HF members across work experience categories of 1-5 years and more than 5 years of experience. While the performance of all HF members with less than one year of experience was comparable. However, the comparison between >35 years and work experience could not be interpreted due to the very small number of participants.

### Level 4: System-level impact

At Tier 2, individual-level assessments of psychosocial stressors, MHPSS needs, duration of mental health condition and assessment of social support provided a community-level profile. The outcome-based data (interventions & referrals) provided system-level insights about the effectiveness of the tiered model, functional referral pathway, as well as the broader resource and service requirements needed to strengthen the system.

The outcomes of the referred cases, as assessed and managed by the CPs (at tier 2), are presented in Table 5 below:

**Table 5:** Outcome of referred cases (n=549)

| **Table 5: Outcome of referred cases (n=549)** |  |
| --- | --- |
| **Domain/ Variable/Description** | **n (%)** |
| **a. Psychosocial stressors** |  |
| Socioeconomic | 186 (33.9) |
| Family-related | 155 (28.2) |
| Health-related | 133 (24.2) |
| Caregiver burden | 87 (15.8) |
| Education-related | 82 (14.9) |
| Bereavement | 65 (11.8) |
| Relationship | 65 (11.8) |
| Marital | 59 (10.7) |
| Occupational | 50 (9.1) |
| Others | 36 (6.6) |
| Crisis | 25 (4.6) |
| GBV | 21 (3.8) |
| **b. MHPSS needs** |  |
| Mental health: cases where a mental disorder is suspected and clinical or pharmacological intervention is indicated (with or without psychosocial interventions) | 389 (70.9) |
| Psychosocial: cases of stress-related or other conditions where only psychosocial interventions are indicated | 155 (28.2) |
| Medical: cases where medical examination and/or treatment is indicated for physical conditions | 112 (20.4) |
| Economic: cases where financial assistance is indicated as a primary concern | 102 (18.6) |
| Educational/vocational: cases where intervention is indicated to facilitate educational/vocational training | 43 (7.8) |
| Social: cases where concerns related to work or livelihood need to be addressed | 37 (6.7) |
| Others: cases where concerns related to legal, protection etc. need to be addressed | 3 (0.5) |
| **c. Duration of mental health condition** |  |
| Years | 274 (49.9) |
| Months | 118 (21.5) |
| Weeks | 7 (1.3) |
| Not reported | 150 (27.3) |
| **d. Social support** |  |
| Social support present | 344 (62.7) |
| Social support absent | 205 (37.3) |
| **e. Psychosocial interventions** (following mhGAP-HIG guidelines) |  |
| Psychoeducation | 350 (63.8) |
| Slow breathing | 279 (50.8) |
| Basic psychosocial support | 269 (49.0) |
| Strengthening social support | 176 (32.1) |
| Sleep hygiene | 131 (23.9) |
| Promote daily functioning | 95 (17.3) |
| Problem-solving technique | 45 (8.2) |
| Grounding technique | 38 (6.9) |
| Behaviour modification | 12 (2.2) |
| Others | 11 (2.0) |
| Note: 277 cases were also guided to download and use MyCare+ for managing stress |  |
| **f. Referral pathways** |  |
| Primary care physicians | 215 (39.2) |
| MHPSS team | 51 (9.3) |
| Local psychiatric service | 41 (7.5) |
| Legal aid | 5 (0.9) |
| Not referred/reported | 237 (43.1) |

Note: For stressors, MHPSS needs and psychosocial interventions, each case might have multiple entries.

**Table 6:** Summary of evaluation of training outcomes based on KP.

| **Kirkpatrick Level** | **What was measured** | **Source of data/tools** | **Key finding** |
| --- | --- | --- | --- |
| Level 1: Reaction | HF satisfaction and perceived relevance of the training | Post-training feedback survey | High satisfaction with course content, design, and relevance to the cultural and healthcare context. Some challenges with digital literacy |
| Level 2: Learning | Knowledge gain | Pre/post knowledge test | Significant improvement in knowledge |
| Level 3: Behavior & performance | 3.1 Number of active HF | Field performance data and supervision data:<br>3.1 No of HF who referred cases | **3.1** 39.7% of trained HF members were active |
|  | 3.2 Number of valid cases | 3.2 No of cases referred minus no. of invalid cases=no. of valid cases | **3.2** 549 cases (64.2%) were valid |
|  | 3.3 Quality of assessment | 3.3 Comparison of GHQ-12 scores by HF and CP | **3.3** HF had overestimated the psychological distress |
|  | 3.4 Supervisory appraisal | 3.4 Supervisors' ratings & comments | **3.4** Most of HF members were rated as average or above average. |
|  | 3.5 A multi-dimensional evaluation | 3.5 Triangulation of field performance & supervision data | **3.5** Higher supervisor ratings were significantly associated with better field performance |
|  | **3.6** Field performance as a test of recruitment criteria | 3.6 Logistic regression and descriptive analysis | **3.6** Younger age, graduate-level education compared with intermediate education, and <1 year of work experience were independently associated with higher odds of being active. |
| Level 4: System-level Impact | Downstream outcomes following assessment and management by clinical psychologists:<br>4.1 Assessment by CPs<br>a. Psychosocial stressors<br>b. MHPSS needs<br>c. Duration of mental health conditions d. Social support | Dashboards of clinical psychologists/outcome data | **4.1** a. Socioeconomic, family-related, and health-related stressors were the most common identified.<br>b. More than 70% of the cases had mental health needs, followed by psychosocial needs (28.2%) and medical needs (20.4%).<br>c. Almost 50% of the cases had mental health issues lasting for years; and 21% for months.<br>d. 37.3% of the cases had no social support. |
|  | 4.2 Psychosocial interventions |  | **4.2** Most administered psychosocial interventions included psychoeducation (63.8%), slow breathing (50.8%) and basic psychosocial support (49%). |
|  | 4.3 Follow-up and referral pathway<br>a. No of calls<br>b. Further referral |  | **4.3** 39.2% were referred to mhGAP trained primary care physicians and only 7.5% were referred to local psychiatric services. |
| Implementation drivers and barriers |  | FGDs | Inter-sectoral collaboration, online training, ongoing supervision by local supervisors, structured referral pathways, and access to digital tools strengthened the competence of HF and facilitated service delivery.<br>Limited internet connectivity and digital literacy; burden of competing work responsibilities, language constraints, stigma and logistical challenges hindered engagement and performance of HF members. |

### No of calls

For all assigned cases, up to three calls were made to establish contact with the service user as per the protocols. For valid cases, one follow-up session was reported for 131 cases (23.9%), two for 246 cases (44.8%), three for 103 cases (18.8%), and four or more for 28 cases (5.1%). In only 41 cases (7.5%), no follow-up calls were reported.

### Implementation drivers and barriers

**Table 7:**
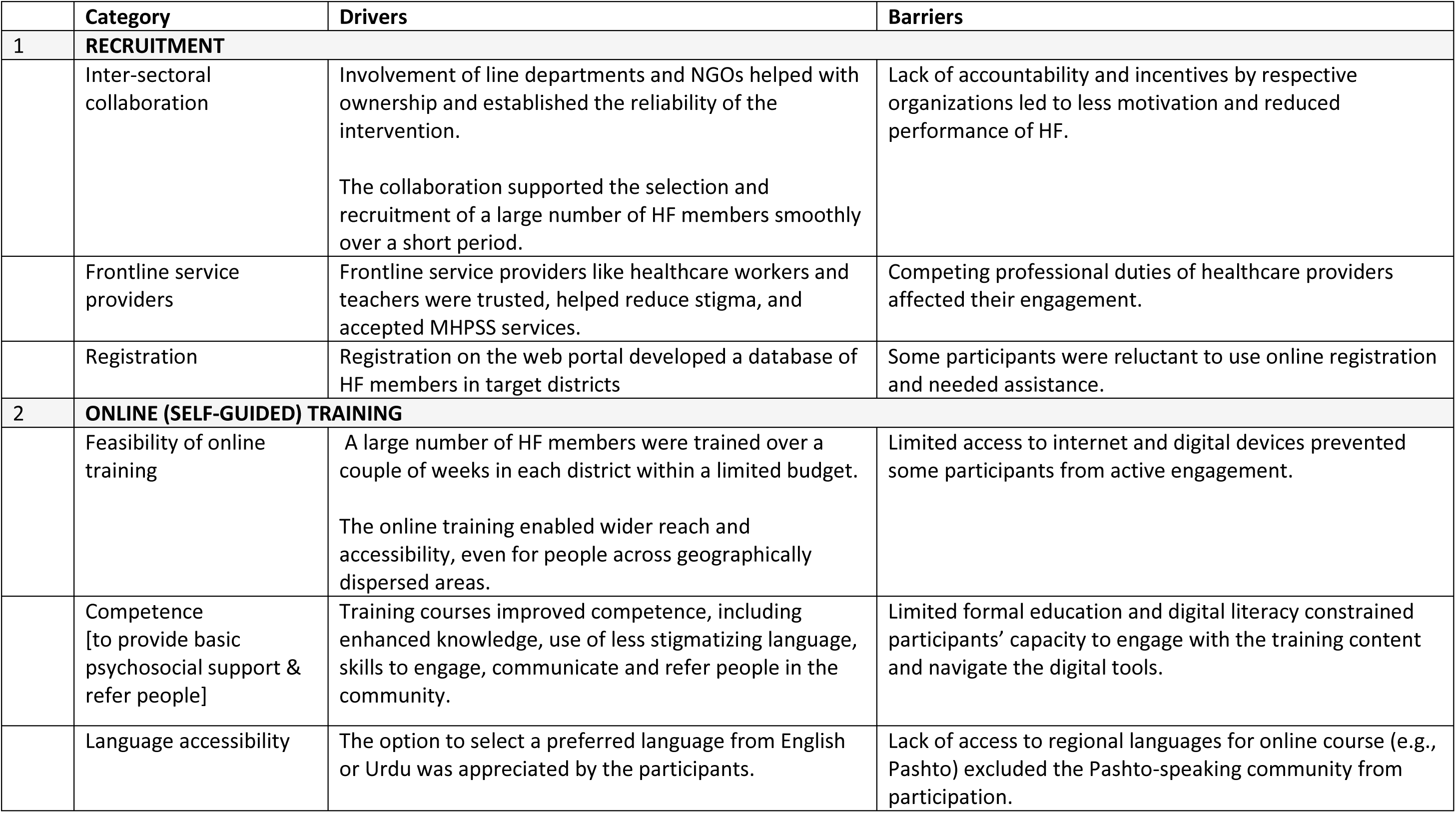
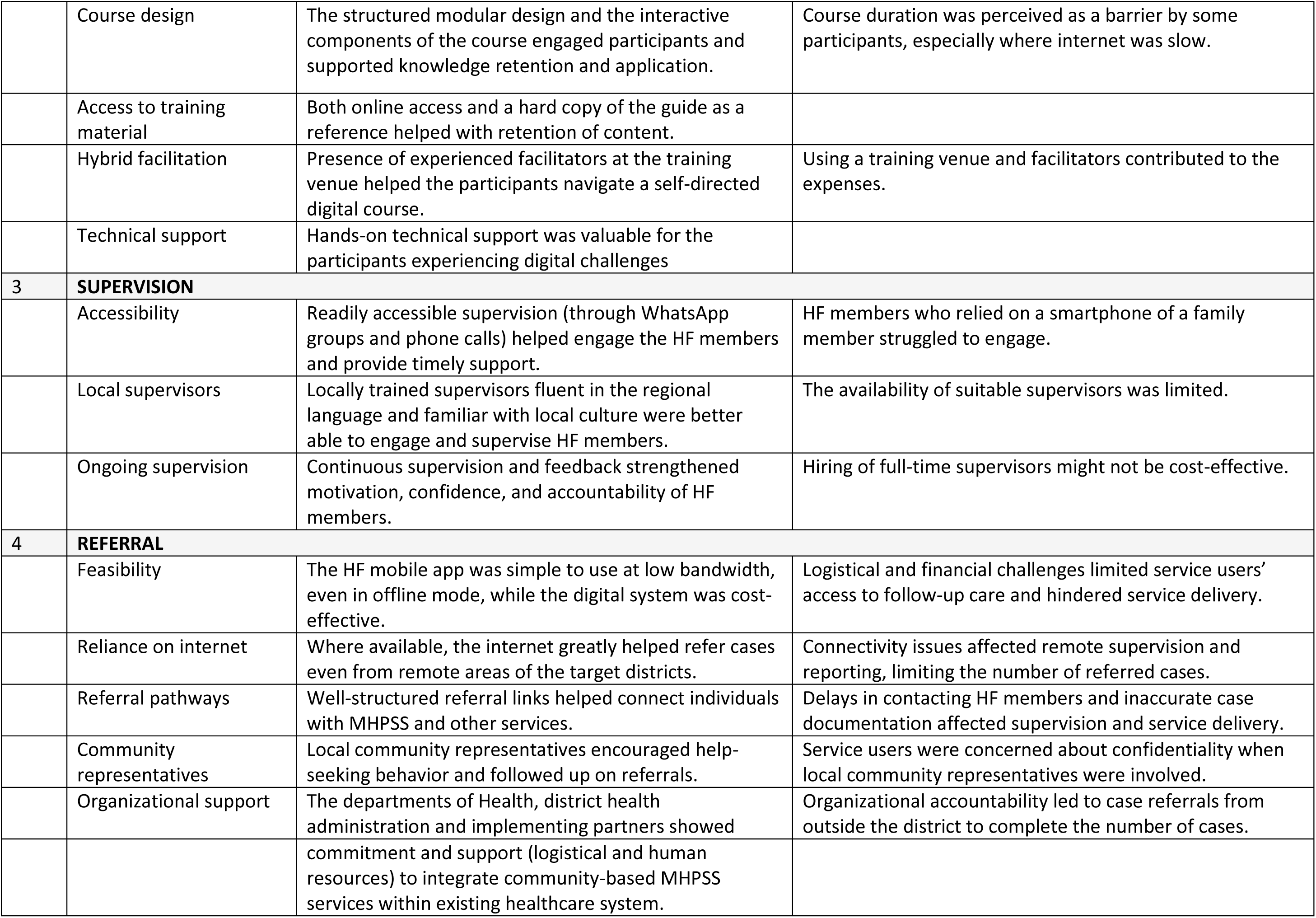
Drivers and barriers of behavioral outcomes.

| Table 7: Drivers and barriers of behavioral outcomes |  |  |  |
| --- | --- | --- | --- |
|  | Category | Drivers | Barriers |
| 1 | **RECRUITMENT** |  |  |
|  | Inter-sectoral collaboration | Involvement of line departments and NGOs helped with ownership and established the reliability of the intervention.<br>The collaboration supported the selection and recruitment of a large number of HF members smoothly over a short period. | Lack of accountability and incentives by respective organizations led to less motivation and reduced performance of HF. |
|  | Frontline service providers | Frontline service providers like healthcare workers and teachers were trusted, helped reduce stigma, and accepted MHPSS services. | Competing professional duties of healthcare providers affected their engagement. |
|  | Registration | Registration on the web portal developed a database of HF members in target districts | Some participants were reluctant to use online registration and needed assistance. |
| 2 | **ONLINE (SELF-GUIDED) TRAINING** |  |  |
|  | Feasibility of online training | A large number of HF members were trained over a couple of weeks in each district within a limited budget.<br>The online training enabled wider reach and accessibility, even for people across geographically dispersed areas. | Limited access to internet and digital devices prevented some participants from active engagement. |
|  | Competence [to provide basic psychosocial support & refer people] | Training courses improved competence, including enhanced knowledge, use of less stigmatizing language, skills to engage, communicate and refer people in the community. | Limited formal education and digital literacy constrained participants' capacity to engage with the training content and navigate the digital tools. |
|  | Language accessibility | The option to select a preferred language from English or Urdu was appreciated by the participants. | Lack of access to regional languages for online course (e.g., Pashto) excluded the Pashto-speaking community from participation. |
|  | Course design | The structured modular design and the interactive components of the course engaged participants and supported knowledge retention and application. | Course duration was perceived as a barrier by some participants, especially where internet was slow. |
|  | Access to training material | Both online access and a hard copy of the guide as a reference helped with retention of content. |  |
|  | Hybrid facilitation | Presence of experienced facilitators at the training venue helped the participants navigate a self-directed digital course. | Using a training venue and facilitators contributed to the expenses. |
|  | Technical support | Hands-on technical support was valuable for the participants experiencing digital challenges |  |
| **3** | **SUPERVISION** |  |  |
|  | Accessibility | Readily accessible supervision (through WhatsApp groups and phone calls) helped engage the HF members and provide timely support. | HF members who relied on a smartphone of a family member struggled to engage. |
|  | Local supervisors | Locally trained supervisors fluent in the regional language and familiar with local culture were better able to engage and supervise HF members. | The availability of suitable supervisors was limited. |
|  | Ongoing supervision | Continuous supervision and feedback strengthened motivation, confidence, and accountability of HF members. | Hiring of full-time supervisors might not be cost-effective. |
| **4** | **REFERRAL** |  |  |
|  | Feasibility | The HF mobile app was simple to use at low bandwidth, even in offline mode, while the digital system was cost-effective. | Logistical and financial challenges limited service users' access to follow-up care and hindered service delivery. |
|  | Reliance on internet | Where available, the internet greatly helped refer cases even from remote areas of the target districts. | Connectivity issues affected remote supervision and reporting, limiting the number of referred cases. |
|  | Referral pathways | Well-structured referral links helped connect individuals with MHPSS and other services. | Delays in contacting HF members and inaccurate case documentation affected supervision and service delivery. |
|  | Community representatives | Local community representatives encouraged help-seeking behavior and followed up on referrals. | Service users were concerned about confidentiality when local community representatives were involved. |
|  | Organizational support | The departments of Health, district health administration and implementing partners showed commitment and support (logistical and human resources) to integrate community-based MHPSS services within existing healthcare system. | Organizational accountability led to case referrals from outside the district to complete the number of cases. |

## Discussion

This study evaluated the training, performance and system-level outcomes of the Hamdard Force intervention, a culturally adapted, digital community-based intervention in the humanitarian settings of Pakistan (Humayun et al., 2025b). We applied the Kirkpatrick framework for a structured, multi-level evaluation of the HF intervention, starting from participant reaction and learning to their behavior and system-level impact using a mixed-methods approach (Kirkpatrick & Kirkpatrick, 2016). We also identified contextual factors that influenced the implementation.

The objectives for HF align with the broader recommendations in the literature, as the intervention is designed to bridge the gap in access to mental health services through early detection at the community level, refer cases using structured referral pathways, and combat prevailing stigma related to mental disorders (Castillo et al., 2019; Jordans et al., 2015). We also offer a chain of evidence, recognizing critical components for sustained practice, including ongoing supervision, monitoring and evaluation mechanisms, quality assurance and organizational support, as highlighted in the literature (Kirkpatrick & Kirkpatrick, 2016; Murray et al., 2011; Singla et al., 2017).

We tried to address the implementation challenges identified with community mental health interventions in LMICs, which underscore the need for locally adapted, stakeholder-driven implementation strategies (Giebel et al., 2024). Rather than a stand-alone training, the intervention serves as the entry point to an integrated MHPSS system. Features like recruitment of community members through intersectoral collaboration; their registration on a digital platform; offering them hybrid training and supervision using contextualized protocols; and connecting them through a mobile application to the MHPSS digital portal; support it as a rights-based, scalable intervention (Humayun et al., 2025b).

The NWKM evaluation of the HF training provided insights beyond knowledge acquisition and helped to examine participants’ reactions, learning, behavior change and implementation outcomes across four interconnected levels (Kirkpatrick & Kirkpatrick, 2016). Level 1 findings indicate cultural relevance and acceptance of the intervention, reinforcing the importance of designing culturally responsive community mental health training programs (Haroz et al., 2020; Upadhyay et al., 2023); more likely to be successfully adopted and integrated into routine practice (Damschroder et al., 2022; Proctor et al., 2011). The acceptability and flexibility of the e-learning approach also support the feasibility of technology-assisted training of community mental health workers, especially in resource-constrained settings (Naslund et al., 2017).

At level 2 (Learning), the training course led to significant improvements in the knowledge and confidence of HF members, and ongoing structured supervision helped motivate and guide them to identify and refer individuals with mental health needs, indicating strengthening of competencies. Our experience strongly suggests that specialist-led supervision is critical to ensure the commitment of the community mental health workforce (Bolton et al., 2023). In our case, re-assessment of screening for psychological well-being performed by the HF members also reinforces the need for ongoing supervision, as the HF overestimated the severity of distress and rated much higher levels of GHQ scores. We had selected local specialists as supervisors, who were familiar with cultural norms, practices, and field-related challenges, and contributed to effective implementation of the intervention (Patel et al., 2013; World Health Organization, 2021).

An early system-level impact is evident through the behavior change at Level 3, i.e., identification and referral of people with MHPSS needs. The level of behavior transfer is consistent with other reported community mental health interventions, which concluded that implementation outcomes remain dependent on continuous supervision, monitoring, access to resources, stigma, and reliable mental health systems (Abas et al., 2016; Patena et al., 2025).

The evaluation of the performance of HF revealed that younger HF members, particularly those with graduate-level of education and less than one year of work experience, were more active in the field. The performance of younger HF members can be attributed to their greater willingness and receptivity to newly developed training programs and adaptability to new learning approaches such as digital learning platforms (Patel et al., 2013; Singla et al., 2017). Higher levels of mental health literacy and confidence in younger members are also known to enable them to implement referral guidelines more effectively (Hadlaczky et al., 2014; Jorm, 2012). We also learnt that implementation is more successful when the workforce is carefully selected, trained, supervised, and monitored during implementation (Agudelo-Hernández et al., 2025).

We were cognizant that any community mental health intervention is less likely to be effective unless it is supported by a broader mental healthcare system and an effective referral mechanism. Our MHPSS service model, based on Inter-Agency Standing Committee guidelines (IASC, 2007) offers a tiered approach where all cases reported by HF at Tier 1 were triaged and managed by CPs who provided psychosocial interventions at Tier 2. It is well established that despite evidence of the efficacy and effectiveness of psychosocial and pharmacological interventions, access to these treatments remains critically insufficient in LMICs (Patel et al., 2018). Approximately a third of reported cases, where pharmacological interventions were indicated, were referred to the nearest trained primary care physicians at Tier 3 of our model. Consistent with Patel et al (2018), our service model is likely to reduce the burden on scarce specialist services locally, as only a small proportion of cases required referral to psychiatric services. At level 4, these findings helped evaluate the system-level impact of the HF intervention.

The triangulation approach enriched the results of our study beyond a single data source, offering a nuanced understanding of the implementation process. Consistent themes across data sources emerged, underscoring the importance of intersectoral collaboration, contextually adapted training, consistent supervision, and functional referral pathways. The intersectoral collaboration facilitated participant referrals, reduced stigma in help-seeking behaviors and developed a database of HF members in target districts. Due to a lack of organizational accountability and incentives, and the burden of competing professional duties, the field performance of HF was affected.

Despite evidence that mobile technologies contribute to optimizing task-shifting for community-based mental healthcare delivery, limitations of digital infrastructure have also been reported in resource-limited settings (Chakrabarti, 2024). The HF intervention has been designed to address the challenges related to internet access or the mobile application. In such cases, the HF members can directly refer cases to their supervisors (CPs) through a phone call or text message.

## Limitations and future directions

The pilot implementation of the HF intervention was the first step towards setting up a system for community mental healthcare in the province of Khyber Pakhtunkhwa; and collecting data atthe grassroots level. We were not able to measure the medium- or long-term impact on community awareness, reduction of stigma, or any changes in help-seeking behavior. A follow-up evaluation at 12 months is recommended to assess broader Level 4 outcomes. In the longer term, it would be crucial to set target indicators for improved mental health and psychosocial wellbeing of community members, such as early identification of mental health conditions and impact of the service, such as effectiveness of referral pathways.

We employed purposive sampling to recruit the participants, which is likely to introduce selection bias, such as overrepresentation of females in our study due to the selection of lady health workers by the health department. Based on the results of this study, future research may revise the recruitment criteria and consider probability-based sampling to remove selection bias.

We observed limited post-training field activity, which may be addressed by engaging the partner organizations effectively and incentivizing the workforce. Overall, the transfer of training into routine practice was partial yet encouraging, with evidence of meaningful changes in referral behavior despite significant contextual challenges limiting implementation fidelity.

## Authors Contribution

AH: Conceptualization, Methodology, Visualization, Supervision, Project administration, Writing – review and editing.

NM: Writing – original draft, data curation, project implementation. AN: Methodology, formal analysis.

AA: Writing – original draft.

HH: Project administration, Supervision

## Data Availability

The data that support the findings of this study are available from the Health Section at the Ministry of Planning, Development & Special Initiatives, Government of Pakistan. Confidentiality restrictions apply to the availability of the data used under the license for the current study, and so they are not publicly available. However, the data can be made available from the authors upon reasonable request after formal approval from the Ministry of Planning, Development & Special Initiatives, Pakistan.

## Acknowledgments

The implementation of the pilot project was supported by the German Federal Ministry of Economic Cooperation and Development (BMZ) through its implementing agency Deutsche Gesellschaft für Internationale Zusammenarbeit (GIZ) GmbH. As grant partners of GIZ, International Medical Corps (IMC) and Handicap International (HI) also supported the implementation.

We would like to acknowledge the valuable contributions of clinical psychologists Adnan Raza, Sohaib Khan, and Memoona Nadir from IMC; clinical psychologists Naveeda Khattak and Sadia Saif from HI for providing services; and Dr. Arooj Najmussaqib for supervising the clinical psychologists at Tier 2.

## Ethical Statement

Ethical approval for the study was provided by the Health Section, Ministry of Planning, Development and Special Initiatives, Government of Pakistan, under letter No. 6 (262) HPC/2026. The study was conducted under the oversight of the Health Section to ensure that MHPSS monitoring, evaluation, and research activities followed relevant ethical standards and regulatory requirements.

Informed verbal consent was obtained from all the participants. Our study adheres to the Declaration of Helsinki, and the reported data comply with the declaration (World Medical Association, 2013).

## List of abbreviations

HF: Hamdard Force
CPs: Clinical Psychologists
PFA: Psychological First Aid
MHPSS: Mental Health and Psychosocial Support
MoPD&SI: Ministry of Planning, Development and Special Initiatives
MHSP&C Unit: Mental Health Strategic Planning and Coordination Unit
FGDs: Focus Group Discussions
GHQ-12: 12-item General Health Questionnaire
LMICs: Low-and-middle-income countries
LMS: Learning Management System
NWKM: New World Kirkpatrick Model.

## Supplementary material

### Supplementary Material A

Questions from MHPSS team:

1. How was your experience coordinating with other departments/organizations?
2. How was your experience regarding the selection of the participants/their recruitment?
3. How was your experience in conducting trainings?
4. What were the barriers to the trainings/implementation?
5. What were the facilitators during implementation?
6. How can this intervention be optimized in the future?
7. What would help make the program more effective and sustainable, considering the context of KP, particularly Haripur and Kohat?

Questions from HF supervisors:

1. Did the participants experience any difficulty in the training, particularly related to the digital platform?
2. What were the barriers to the trainings/implementation?
3. What were the facilitators during implementation?
4. How can this intervention be optimized in the future?
5. How effective was the contribution/role of the HF members in facilitating the communication between service provider (CP) and the service user (person with MH needs)?
6. Were there any challenges identified related to HF members’ supervision/communication?
7. What would help make the program more effective and sustainable considering the context of KP?

### Supplementary material B

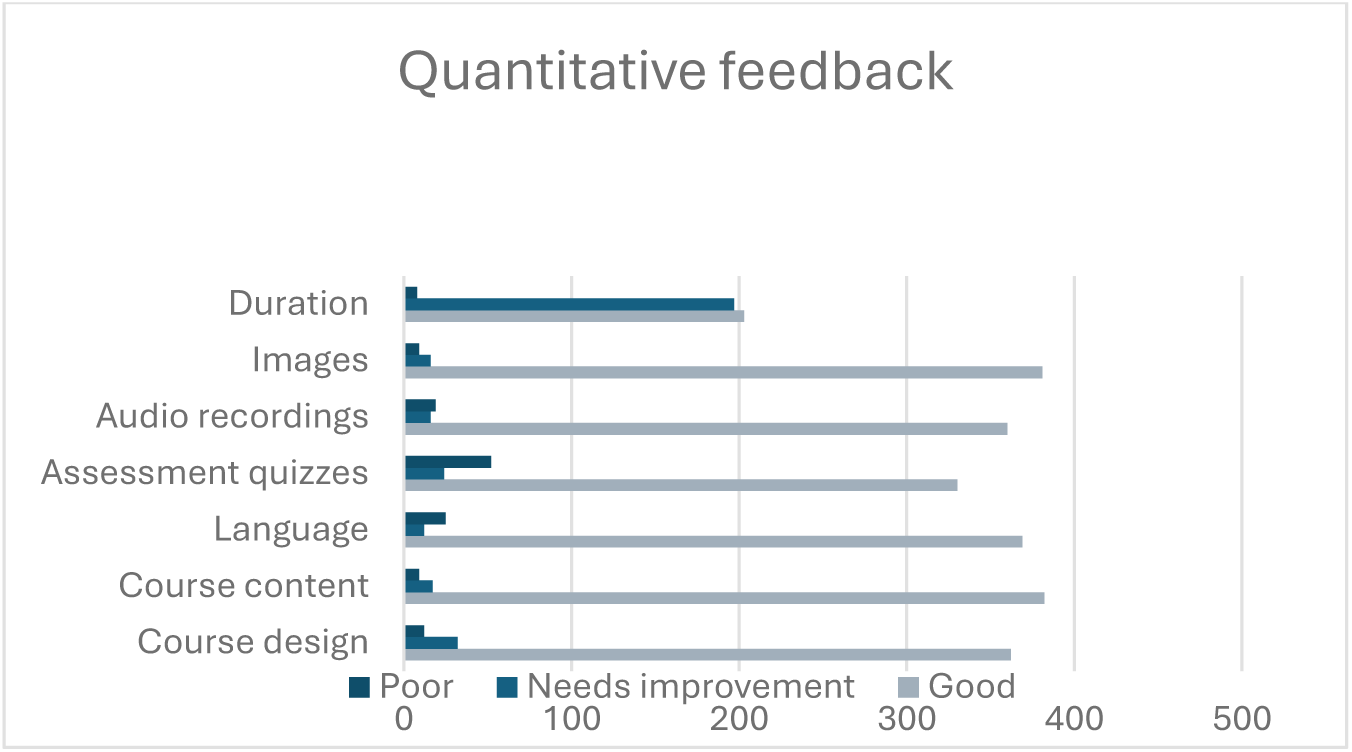

### Supplementary material C

**Table S2.**
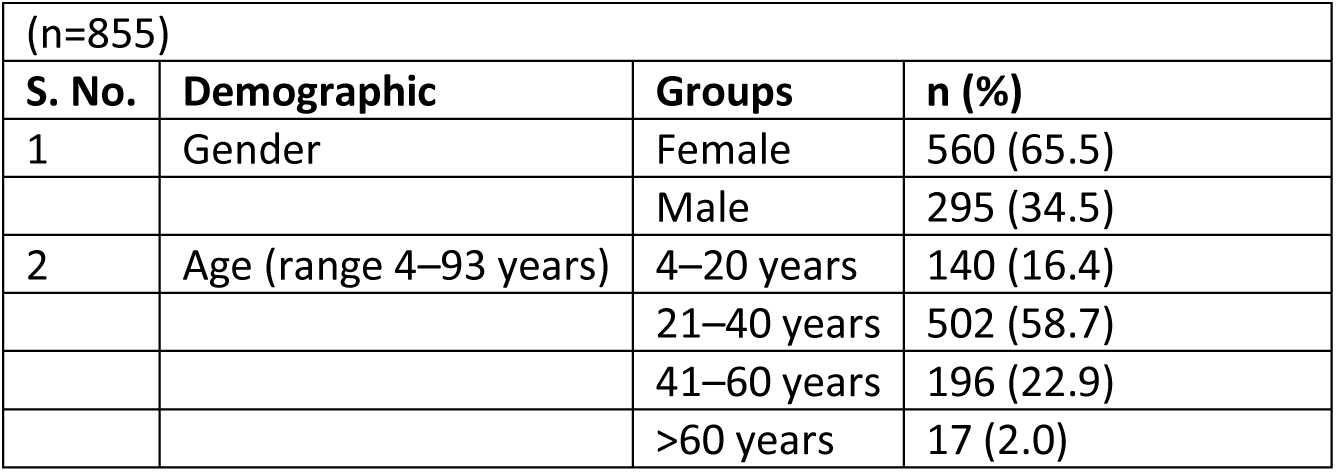
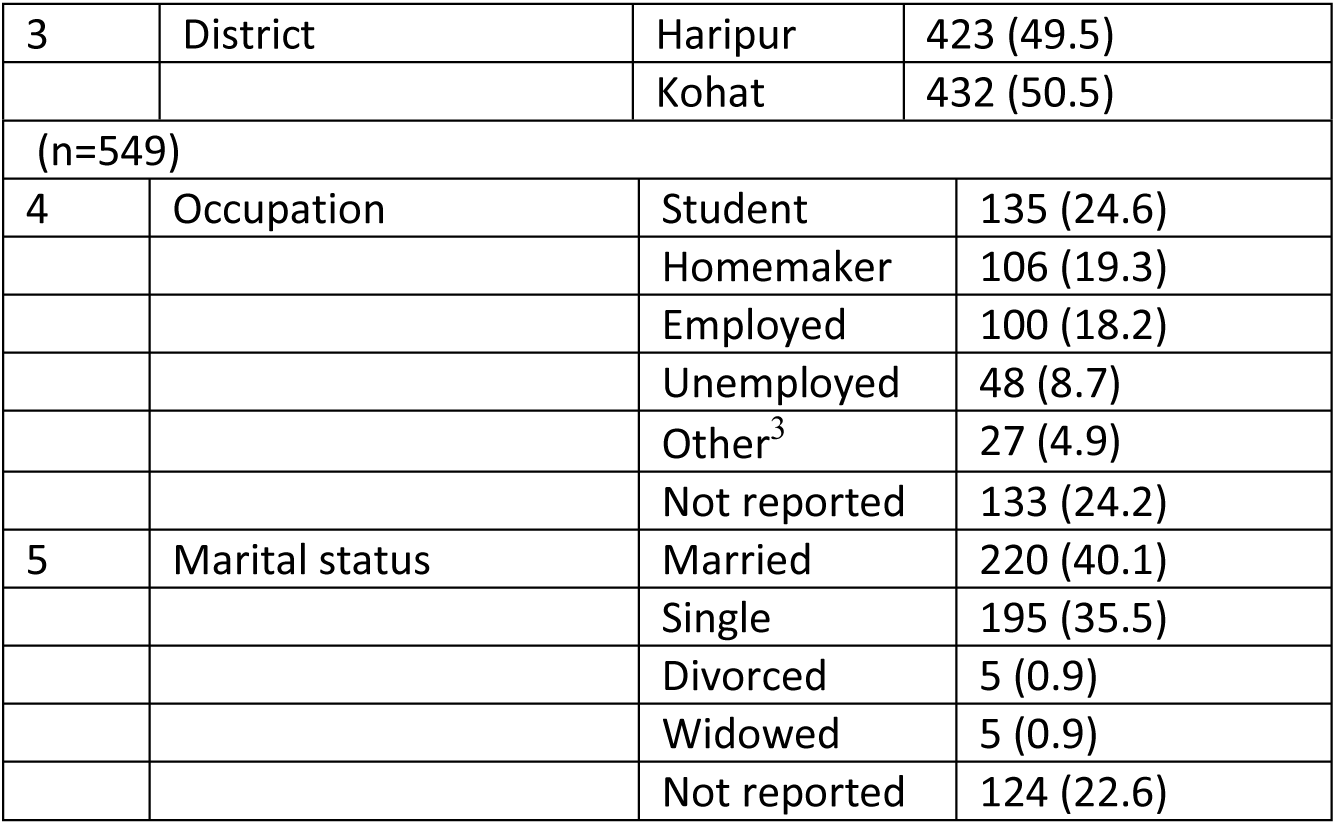
Sociodemographic characteristics of the referred cases.

### Supplementary material D

**Table S3.** Cross-tabulation of case submission rates by age, education level, and work Experience.

|  |  |  |  |
| --- | --- | --- | --- |
| **Participant characteristic** | **Category** | **≤35 years n/N (%)** | **>35 years n/N (%)** |
| Education level | Intermediate | 14/39 (35.9) | 25/122 (20.5) |
|  | Graduation | 48/88 (54.5) | 17/41 (41.5) |
|  | Master's or above | 40/63 (63.5) | 16/50 (32.0) |
| Work experience | <1 year | 35/48 (72.9) | 3/4 (75.0) |
|  | 1–5 years | 32/57 (56.1) | 2/14 (14.3) |
|  | >5 years | 35/85 (41.2) | 53/195 (27.2) |

Note: $n$ = number of HF members who submitted at least one case; $N$ = total number of HF members in the corresponding subgroup. Percentages represent case submission rates ( $n/N \times 100$ ).

## Footnotes

1 https://mhpss.pk/

3 Other includes retired, daily wage workers, self-employed, and other occupations

## References

Abas, M., Bowers, T., Manda, E., Cooper, S., Machando, D., Verhey, R., Lamech, N., Araya, R., & Chibanda, D. (2016). ‘Opening up the mind’: problem-solving therapy delivered by female lay health workers to improve access to evidence-based care for depression and other common mental disorders through the Friendship Bench Project in Zimbabwe. International Journal of Mental Health Systems, 10(1), 39. 10.1186/s13033-016-0071-9

Agudelo-Hernández, F., Vélez-Botero, H., Plata-Casas, L. I., & Delgado-Reyes, A. C. (2025). Factors driving implementation success of mental health gap action programme (mhGAP) in Colombia: a cross-sectional study. Discover Mental Health, 5(1), 152. 10.1007/s44192-025-00282-7

Bolton, P., West, J., Whitney, C., Jordans, M. J. D., Bass, J., Thornicroft, G., Murray, L., Snider, L., Eaton, J., Collins, P. Y., Ventevogel, P., Smith, S., Stein, D. J., Petersen, I., Silove, D., Ugo, V., Mahoney, J., el Chammay, R., Contreras, C., … Raviola, G. (2023). Expanding mental health services in low- and middle-income countries: A task-shifting framework for delivery of comprehensive, collaborative, and community-based care. Cambridge Prisms: Global Mental Health, 10, e16. 10.1017/gmh.2023.5

Castillo, E. G., Ijadi-Maghsoodi, R., Shadravan, S., Moore, E., Mensah, M. O., Docherty, M., Aguilera Nunez, M. G., Barcelo, N., Goodsmith, N., Halpin, L. E., Morton, I., Mango, J., Montero, A. E., Rahmanian Koushkaki, S., Bromley, E., Chung, B., Jones, F., Gabrielian, S., Gelberg, L., … Wells, K. B. (2019). Community Interventions to Promote Mental Health and Social Equity. Current Psychiatry Reports, 21(5), 35. 10.1007/s11920-019-1017-0

Chakrabarti, S. (2024). Digital psychiatry in low-and-middle-income countries: New developments and the way forward. World Journal of Psychiatry, 14(3), 350–361. 10.5498/wjp.v14.i3.350

Damschroder, L. J., Reardon, C. M., Widerquist, M. A. O., & Lowery, J. (2022). The updated Consolidated Framework for Implementation Research based on user feedback. Implementation Science, 17(1), 75. 10.1186/s13012-022-01245-0

Fox, J. H., Burkle, F. M., Bass, J., Pia, F. A., Epstein, J. L., & Markenson, D. (2012). The Effectiveness of Psychological First Aid as a Disaster Intervention Tool: Research Analysis of Peer-Reviewed Literature From 1990-2010. Disaster Medicine and Public Health Preparedness, 6(3), 247–252. 10.1001/dmp.2012.39

Giebel, C., Gabbay, M., Shrestha, N., Saldarriaga, G., Reilly, S., White, R., Liu, G., Allen, D., & Zuluaga, M. I. (2024). Community-based mental health interventions in low- and middle-income countries: a qualitative study with international experts. International Journal for Equity in Health, 23(1), 19. 10.1186/s12939-024-02106-6

Global Burden of Disease. (2022). Global, regional, and national burden of 12 mental disorders in 204 countries and territories, 1990–2019: a systematic analysis for the Global Burden of Disease Study 2019. The Lancet Psychiatry, 9(2), 137–150. 10.1016/S2215-0366(21)00395-3

Hadlaczky, G., Hökby, S., Mkrtchian, A., Carli, V., & Wasserman, D. (2014). Mental Health First Aid is an effective public health intervention for improving knowledge, attitudes, and behaviour: A meta-analysis. International Review of Psychiatry, 26(4), 467–475. 10.3109/09540261.2014.924910

Haroz, E., Nguyen, A., Lee, C., Tol, W., Fine, S., & Bolton, P. (2020). What works in psychosocial programming in humanitarian contexts in low- and middle-income countries: a systematic review of the evidence. Intervention, 18(1), 3. 10.4103/INTV.INTV_6_19

Humayun, A. (2021). A rights-based digital solution for public mental healthcare in Pakistan. Ministry of Planning, Development and Special Initiatives, Government of Pakistan. https://www.pc.gov.pk/web/blog/getblog/14

Humayun, A., Haq, I., Khan, F., & Nasir, S. (2020). Designing Psychosocial Support for COVID-19 Frontline Responders in Pakistan: A Potentially Scalable Self-Help Plus Blueprint for LMICs. Intervention, 18(2), 150. 10.4103/INTV.INTV_21_20

Humayun, A., Najmussaqib, A., & Muneeb, N. ul A. (2025a). Developing and feasibility testing of Hamdard Force: an intervention to build the capacity of a community mental health workforce in Pakistan. BMC Public Health. 10.1186/s12889-025-25701-w

Humayun, A., Najmussaqib, A., & Muneeb, N. ul A. (2025b). Developing and feasibility testing of Hamdard Force: an intervention to build the capacity of a community mental health workforce in Pakistan. BMC Public Health, 26(1), 198. 10.1186/s12889-025-25701-w

IASC. (2007). Inter-Agency Standing Committee guidelines on Mental Health and Psychosocial Support in Emergency Settings. https://interagencystandingcommittee.org/sites/default/files/migrated/2020-11/IASC Guidelines on Mental Health and Psychosocial Support in Emergency Settings %28English%29.pdf

Jordans, M. J. D., Kohrt, B. A., Luitel, N. P., Komproe, I. H., & Lund, C. (2015). Accuracy of proactive case finding for mental disorders by community informants in Nepal. British Journal of Psychiatry, 207(6), 501–506. 10.1192/bjp.bp.113.141077

Jorm, A. F. (2012). Mental health literacy: Empowering the community to take action for better mental health. American Psychologist, 67(3), 231–243. 10.1037/a0025957

Kirkpatrick, J. D., & Kirkpatrick, W. K. (2016). Kirkpatrick’s four levels of training evaluation. Association for Talent Development (ATD) Press. Association for Talent Development (ATD) Press. https://books.google.com.pk/books?hl=en&lr=&id=mo--DAAAQBAJ&oi=fnd&pg=PT10&dq=Kirkpatrick,+J.+D.,+%26+Kirkpatrick,+W.+K.+(2016).+Kirkpatrick%27s+four+levels+of+training+evaluation.+Association+for+Talent+Development+(ATD)+Press.&ots=LOKcRLkiQC&sig=rgCXwhDEI5DNLKOy6khQgUJWFmE#v=onepage&q&f=false

Liao, S.-C., & Hsu, S.-Y. (2019). Evaluating A Continuing Medical Education Program: New World Kirkpatrick Model Approach. *International Journal of Management*, Economics and Social Sciences, 8(4). 10.32327/IJMESS/8.4.2019.17

Moitra, M., Santomauro, D., Collins, P. Y., Vos, T., Whiteford, H., Saxena, S., & Ferrari, A. J. (2022). The global gap in treatment coverage for major depressive disorder in 84 countries from 2000– 2019: A systematic review and Bayesian meta-regression analysis. PLOS Medicine, 19(2), e1003901. 10.1371/journal.pmed.1003901

MoPD&SI. (2025). Mental health and psychosocial support (MHPSS) model. Mental Health Strategic Planning & Coordination Unit. Ministry of Planning, Development and Special Initiatives, Islamabad. https://mhpss.pk/mhpss-model

Morgan, H. (2024). Using Triangulation and Crystallization to Make Qualitative Studies Trustworthy and Rigorous. The Qualitative Report. 10.46743/2160-3715/2024.6071

Murray, L. K., Dorsey, S., Bolton, P., Jordans, M. J., Rahman, A., Bass, J., & Verdeli, H. (2011). Building capacity in mental health interventions in low resource countries: an apprenticeship model for training local providers. International Journal of Mental Health Systems, 5(1), 30. 10.1186/1752-4458-5-30

Naslund, J. A., Aschbrenner, K. A., Araya, R., Marsch, L. A., Unützer, J., Patel, V., & Bartels, S. J. (2017). Digital technology for treating and preventing mental disorders in low-income and middle-income countries: a narrative review of the literature. The Lancet Psychiatry, 4(6), 486–500. 10.1016/S2215-0366(17)30096-2

Naveed, S., Waqas, A., Chaudhary, A. M. D., Kumar, S., Abbas, N., Amin, R., Jamil, N., & Saleem, S. (2020). Prevalence of Common Mental Disorders in South Asia: A Systematic Review and Meta-Regression Analysis. Frontiers in Psychiatry, 11. 10.3389/fpsyt.2020.573150

Patel, V., Belkin, G. S., Chockalingam, A., Cooper, J., Saxena, S., & Unützer, J. (2013). Grand Challenges: Integrating Mental Health Services into Priority Health Care Platforms. PLoS Medicine, 10(5), e1001448. 10.1371/journal.pmed.1001448

Patel, V., Saxena, S., Lund, C., Thornicroft, G., Baingana, F., Bolton, P., Chisholm, D., Collins, P. Y., Cooper, J. L., Eaton, J., Herrman, H., Herzallah, M. M., Huang, Y., Jordans, M. J. D., Kleinman, A., Medina-Mora, M. E., Morgan, E., Niaz, U., Omigbodun, O., … UnÜtzer, Jü. (2018). The Lancet Commission on global mental health and sustainable development. The Lancet, 392(10157), 1553–1598. 10.1016/S0140-6736(18)31612-X

Patel, V., Saxena, S., Lund, C., Thornicroft, G., &, et al. (2018). The Lancet Commission on global mental health and sustainable development. The Lancet, 392(10157), 1553–1598. https://www.thelancet.com/journals/lancet/article/PIIS0140-6736(18)31612-X/abstract

Patena, J., Adenikinju, D., Lanka, P., Hameed, T., Kulkarni, S., Osei-Tutu, N., Zuniga, S., Ruan, C., Shenoy, S., Thakkar, D., Noble, E., Angulo, B., Vieira, D., Gyamfi, J., & Peprah, E. (2025). Evaluating implementation research outcomes for a task-sharing mental health intervention: A systematic review of the Friendship Bench. Cambridge Prisms: Global Mental Health, 12, e65. 10.1017/gmh.2025.10025

Perion, J., Steiner, V., Kinney, J., McBride, K., & Saltzman, B. (2021). Applying the New World Kirkpatrick Model to evaluate an online training course introducing a multi-sensory device. Innovation in Aging, 5(Supplement_1), 763–763. 10.1093/geroni/igab046.2827

Proctor, E., Silmere, H., Raghavan, R., Hovmand, P., Aarons, G., Bunger, A., Griffey, R., & Hensley, M. (2011). Outcomes for Implementation Research: Conceptual Distinctions, Measurement Challenges, and Research Agenda. Administration and Policy in Mental Health and Mental Health Services Research, 38(2), 65–76. 10.1007/s10488-010-0319-7

Shultz, J. M., & Forbes, D. (2014). Psychological First Aid. Disaster Health, 2(1), 3–12. 10.4161/dish.26006

Singla, D. R., Kohrt, B. A., Murray, L. K., Anand, A., Chorpita, B. F., & Patel, V. (2017). Psychological Treatments for the World: Lessons from Low- and Middle-Income Countries. Annual Review of Clinical Psychology, 13(1), 149–181. 10.1146/annurev-clinpsy-032816-045217

Upadhyay, K., Goel, S., & John, P. (2023). Developing a capacity building training model for public health managers of low and middle income countries. PLOS ONE, 18(4), e0272793. 10.1371/journal.pone.0272793

World Health Organization. (2021). Guidance on community mental health services: Promoting person-centred and rights-based approaches. https://www.who.int/publications/i/item/9789240025707

World Health Organization. (2022). World mental health report: Transforming mental health for all. https://www.who.int/publications/i/item/9789240049338

World Health Organization, G. (2011). World Health Organization, War Trauma Foundation and World Vision International. Psychological First Aid: Guide for Field Workers.

